# HLA-B*58:01 is an Incomplete Predictor of Allopurinol-Induced Severe Cutaneous Adverse Reactions

**DOI:** 10.1101/2025.05.23.25328236

**Authors:** Chelsea N. Campbell, Matthew S. Krantz, Alexis Yu, Elizabeth J. Phillips, the SJS Survivor Study

## Abstract

**Importance:** Carriage of HLA-B*58:01 has been shown to have a strong association with the development of allopurinol-induced Stevens-Johnson syndrome and toxic epidermal necrolysis (SJS/TEN) and drug reaction and eosinophilia and systemic symptoms (DRESS) in many populations globally; however, there is a critical need to determine if this is generalizable to varying populations including those in the United States (US).

**Objective:** To perform HLA class I and II association studies in a cohort of US patients diagnosed with allopurinol-induced SJS/TEN or DRESS compared to allopurinol tolerant and population controls.

**Design, Setting, and Participants:** We enrolled consenting individuals who had specialist adjudicated allopurinol-induced SJS/TEN or DRESS (collectively allopurinol-SCAR). HLA carriage in these cases was compared to allopurinol tolerant and population controls identified through Vanderbilt University Medical Center (VUMC) BioVU, a biobank which includes 94,489 individuals with imputed human leukocyte antigen (HLA) class I and II typing from genotyping array data.

**Main Outcomes and Measures:** We performed HLA class I and II conditional logistic regression case-control analyses between allopurinol-SCAR cases and both population controls and allopurinol tolerant controls matched on age, sex, and self-identified race. We reported odds ratio (OR) and 95% confidence interval (CI) with Bonferroni corrected *P* (*Pc*) <.05.

**Results:** The conditional logistic regression analyses included allopurinol-SCAR cases (n=16) and 10:1 matched allopurinol tolerant controls (n=160). We found two HLA class I alleles independently associated with increased risk of allopurinol-induced SCAR: HLA-B*58:01 (OR 28 [95% CI, 8.6 – 100.6]) and HLA-A*34:02 (OR 20.6 [95% CI, 3.3 – 131.1]). We did not identify any HLA class II alleles meeting the Pc level of significance.

**Conclusions and Relevance:** We found HLA-B*58:01 to be strongly associated with allopurinol-induced SCAR, generalizing findings from previous studies. Additionally, we found HLA-A*34:02 to be a second independent genetic risk factor for allopurinol-SCAR. These findings underscore the need to conduct specific population-based studies that both reproduce known and uncover novel HLA associations in order to reduce harm through contributions to screening, risk stratification, and diagnosis.

**KEY POINTS:** *Question:* Is the association between HLA-B*58:01 and allopurinol-SCAR generalizable to admixed populations in the US or are additional HLA associations involved?

*Findings:* In this HLA association study with 16 patients of primarily self-identified Black race of adjudicated allopurinol-induced SJS/TEN or DRESS, we demonstrate a strong association with the established risk allele, HLA-B*58:01, and, for the first time, identified HLA-A*34:02 as an additional independent risk factor.

*Meaning:* HLA-B*58:01 is absent in more than one-third of our US cohort of allopurinol-SCAR cases suggesting that more comprehensive screening and diagnostic approaches are necessary to prevent additional cases in genetically heterogenous populations.

## INTRODUCTION

Severe cutaneous adverse reactions (SCAR) are T-cell mediated reactions to medications resulting in significant morbidity and mortality globally. SCAR phenotypes include Stevens-Johnson syndrome and toxic epidermal necrolysis (SJS/TEN) and drug reaction with eosinophilia and systemic symptoms (DRESS). One of the most common drugs implicated in SCAR is the uric acid lowering-agent, allopurinol. While carriage of HLA-B*58:01 has been reported to contribute to 100% of allopurinol-induced SCAR (allopurinol-SCAR) cases in Han Chinese populations^1^, this is not true in Northern and Southern European populations where only 55-64% of cases carried HLA-B*58:01^2,3^. Chronic kidney disease (CKD) has also been shown to increase the risk of allopurinol-SCAR due to excessive accumulation of oxypurinol, the renally excreted metabolite of allopurinol^4^. HLA-B*58:01 carriage and prevalence of CKD are likely contributing factors to the risk of developing SJS/TEN being five times higher in Blacks compared to Whites^5,6^. Despite the increased carriage rate of HLA-B*58:01 in US Blacks, HLA associations with allopurinol-SCAR in heterogenous populations in the US have not been defined.

## METHODS

### BioVU Cohorts

Allopurinol tolerant controls were selected from the VUMC BioVU^7^ population based on availability of MEGA^EX^ array typing, self-reported Black or White race, age ≥ 18 years at time of allopurinol treatment, baseline creatine values available in the 30 days prior to allopurinol initiation, ≥ 90 days of allopurinol treatment (drug era length), absence of any International Classification of Diseases (ICD) codes pertaining to an allopurinol adverse reaction, and absence of an allopurinol-specific allergy label. We performed descriptive statistics on the allopurinol tolerant controls including baseline creatinine and presence of ICD codes for CKD and gout. A 10:1 case:control match by age, sex, and self-reported race to allopurinol-SCAR cases was performed through optimal pair matching using the MatchIt R package^8^.

### Allopurinol-SCAR Cases

Consenting patients (n=14) with specialist adjudicated allopurinol-SCAR were prospectively recruited between 2015-2024 under institutional review board (IRB) approval from VUMC IRB (131836, 150754, and 171900). Specialist adjudicated SJS/TEN patients (n=2) were identified cross-sectionally in the SJS survivor research study who had developed SJS/TEN between 2009-2017 and participated in 2020-2024 under VUM IRB approval (191350).

### HLA Typing

HLA class I and II alleles from BioVU participants were imputed from MEGA^EX^ using SNP2HLA to 4 digit resolution^9^. High-resolution typing of HLA-A, HLA-B, HLA-C, HLA-DP, HLA-DQ, and HLA-DR of allopurinol-SCAR cases was performed using sequence-based typing via Illumina MiSeq as previously described^10^.

### Statistical Analyses

The conditional logistic regression of HLA class I and II alleles comparing allopurinol-SCAR cases to 10:1 matched, allopurinol tolerant controls was performed with the MatchIt and MiDAS R packages^11^. HLA class I and II alleles meeting the unadjusted *P*<.05 along with their odds ratio (OR), 95% confidence intervals (CI), and Bonferroni corrected *P* values (*Pc*) were reported. For alleles with *Pc*<.05, we further compared allopurinol-SCAR cases with BioVU population controls, including self-identified Black, self-identified White, and overall populations by Fisher’s exact test. To evaluate for gene-dose effect of HLA-B*58:01, we performed Fisher’s exact test and calculated OR and 95% CI with Haldane’s modification for heterozygous and homozygous genotypes^12^. We performed drug era persistence analyses in the allopurinol tolerant controls (10:1 matched for risk allele carriage by age, gender, and race) for significant alleles to evaluate any differences in reaching 180 days of treatment using the ggsurvfit R package^13^.

## RESULTS

We identified 16 patients diagnosed with allopurinol-SCAR with high-resolution HLA typing (eTable 1) where a total of 11 (69%) cases self-identified as Black and 5 (31%) as White (Table 1). We identified 1,122 allopurinol tolerant controls from BioVU (eFigure 1) and those of self-identified Black race had higher baseline creatinine values (1.76 mg/dL vs 1.46, mg/dL, *P*<.001) and occurrence of ICD codes corresponding to CKD (90% vs 74%, *P*<.001) compared to those of self-identified White race (eTable 2). From the allopurinol tolerant controls, we identified 160 individuals matched (10:1) by age, gender, and race.

**Table 1:**
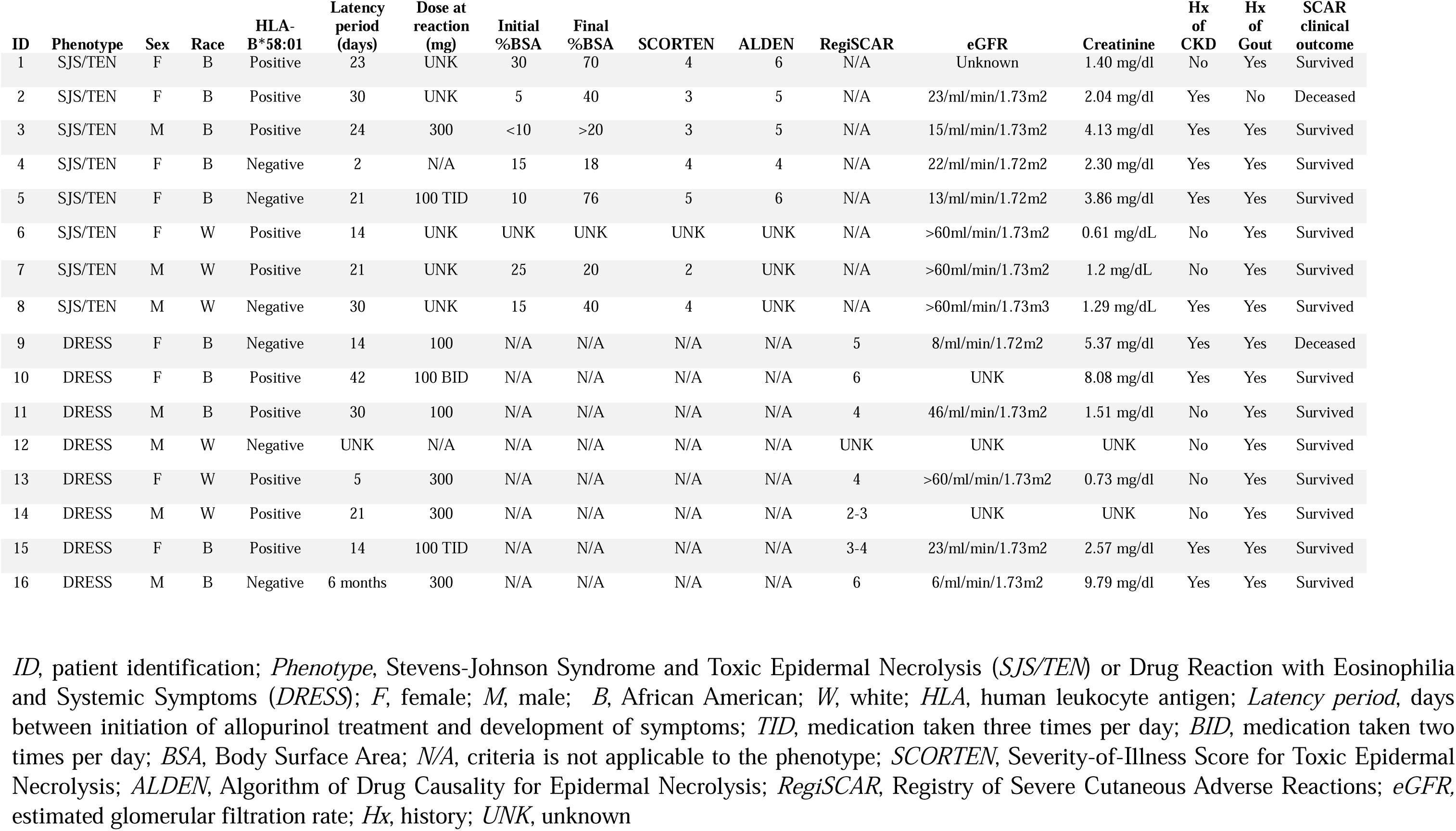
Summary of the allopurinol-associated SCAR case cohort demographics, HLA risk allele carriage, and clinical characteristics.

| ID | Phenotype | Sex | Race | HLA-B*58:01 | Latency period (days) | Dose at reaction (mg) | Initial %BSA | Final %BSA | SCORTEN | ALDEN | RegiSCAR | eGFR | Creatinine | Hx of CKD | Hx of Gout | SCAR clinical outcome |
| --- | --- | --- | --- | --- | --- | --- | --- | --- | --- | --- | --- | --- | --- | --- | --- | --- |
| 1 | SJS/TEN | F | B | Positive | 23 | UNK | 30 | 70 | 4 | 6 | N/A | Unknown | 1.40 mg/dl | No | Yes | Survived |
| 2 | SJS/TEN | F | B | Positive | 30 | UNK | 5 | 40 | 3 | 5 | N/A | 23/ml/min/1.73m2 | 2.04 mg/dl | Yes | No | Deceased |
| 3 | SJS/TEN | M | B | Positive | 24 | 300 | <10 | >20 | 3 | 5 | N/A | 15/ml/min/1.73m2 | 4.13 mg/dl | Yes | Yes | Survived |
| 4 | SJS/TEN | F | B | Negative | 2 | N/A | 15 | 18 | 4 | 4 | N/A | 22/ml/min/1.72m2 | 2.30 mg/dl | Yes | Yes | Survived |
| 5 | SJS/TEN | F | B | Negative | 21 | 100 TID | 10 | 76 | 5 | 6 | N/A | 13/ml/min/1.72m2 | 3.86 mg/dl | Yes | Yes | Survived |
| 6 | SJS/TEN | F | W | Positive | 14 | UNK | UNK | UNK | UNK | UNK | N/A | >60ml/min/1.73m2 | 0.61 mg/dL | No | Yes | Survived |
| 7 | SJS/TEN | M | W | Positive | 21 | UNK | 25 | 20 | 2 | UNK | N/A | >60ml/min/1.73m2 | 1.2 mg/dL | No | Yes | Survived |
| 8 | SJS/TEN | M | W | Negative | 30 | UNK | 15 | 40 | 4 | UNK | N/A | >60ml/min/1.73m3 | 1.29 mg/dL | Yes | Yes | Survived |
| 9 | DRESS | F | B | Negative | 14 | 100 | N/A | N/A | N/A | N/A | 5 | 8/ml/min/1.72m2 | 5.37 mg/dl | Yes | Yes | Deceased |
| 10 | DRESS | F | B | Positive | 42 | 100 BID | N/A | N/A | N/A | N/A | 6 | UNK | 8.08 mg/dl | Yes | Yes | Survived |
| 11 | DRESS | M | B | Positive | 30 | 100 | N/A | N/A | N/A | N/A | 4 | 46/ml/min/1.73m2 | 1.51 mg/dl | No | Yes | Survived |
| 12 | DRESS | M | W | Negative | UNK | N/A | N/A | N/A | N/A | N/A | UNK | UNK | UNK | No | Yes | Survived |
| 13 | DRESS | F | W | Positive | 5 | 300 | N/A | N/A | N/A | N/A | 4 | >60/ml/min/1.73m2 | 0.73 mg/dl | No | Yes | Survived |
| 14 | DRESS | M | W | Positive | 21 | 300 | N/A | N/A | N/A | N/A | 2-3 | UNK | UNK | No | Yes | Survived |
| 15 | DRESS | F | B | Positive | 14 | 100 TID | N/A | N/A | N/A | N/A | 3-4 | 23/ml/min/1.73m2 | 2.57 mg/dl | Yes | Yes | Survived |
| 16 | DRESS | M | B | Negative | 6 months | 300 | N/A | N/A | N/A | N/A | 6 | 6/ml/min/1.73m2 | 9.79 mg/dl | Yes | Yes | Survived |

*ID*, patient identification; *Phenotype*, Stevens-Johnson Syndrome and Toxic Epidermal Necrolysis (*SJS/TEN*) or Drug Reaction with Eosinophilia and Systemic Symptoms (*DRESS*); *F*, female; *M*, male; *B*, African American; *W*, white; *HLA*, human leukocyte antigen; *Latency period*, days between initiation of allopurinol treatment and development of symptoms; *TID*, medication taken three times per day; *BID*, medication taken two times per day; *BSA*, Body Surface Area; *N/A*, criteria is not applicable to the phenotype; *SCORTEN*, Severity-of-Illness Score for Toxic Epidermal Necrolysis; *ALDEN*, Algorithm of Drug Causality for Epidermal Necrolysis; *RegiSCAR*, Registry of Severe Cutaneous Adverse Reactions; *eGFR*, estimated glomerular filtration rate; *Hx*, history; *UNK*, unknown

From our case-control conditional logistic regression analysis, we found two HLA class I alleles independently associated with increased risk of allopurinol-SCAR: HLA-B*58:01 (OR 28 [95% CI, 8.6 – 100.6], *Pc*<.001) and HLA-A*34:02 (OR 20.6 [95% CI, 3.3 – 131.1], *Pc*=.04) (Table 2) while HLA-B*35:01 and HLA-B*58:02 had unadjusted *P*<.05. We did not identify any HLA class II alleles meeting the unadjusted P or Pc levels of significance. Carriage of HLA-B*58:01 (Figure 1A) was higher in the allopurinol-SCAR cases (62.5%) compared to the allopurinol tolerant matched controls (5.6%, *P*<.001) and the BioVU overall population (2.3%, *P*<.001). Carriage of HLA-A*34:02 (Figure 1B) was also higher in the allopurinol-SCAR cases (18.8%) compared to the allopurinol tolerant matched controls (4.4%, *P*=.05) and the BioVU overall population (1.3%, *P*=.001). Homozygosity at HLA-B*58:01 was associated with a higher risk of developing allopurinol-SCAR (OR 55.3 [95% CI, 2.5-1208.6]) compared to heterozygosity in HLA-B*58:01 (eTable 3). Drug era persistence analyses showed no difference in allopurinol tolerant controls with and without HLA-B*58:01 (eFigure 2A) or HLA-A*34:02 reaching a drug era length of 180 days (eFigure 2B).

**Figure 1:**
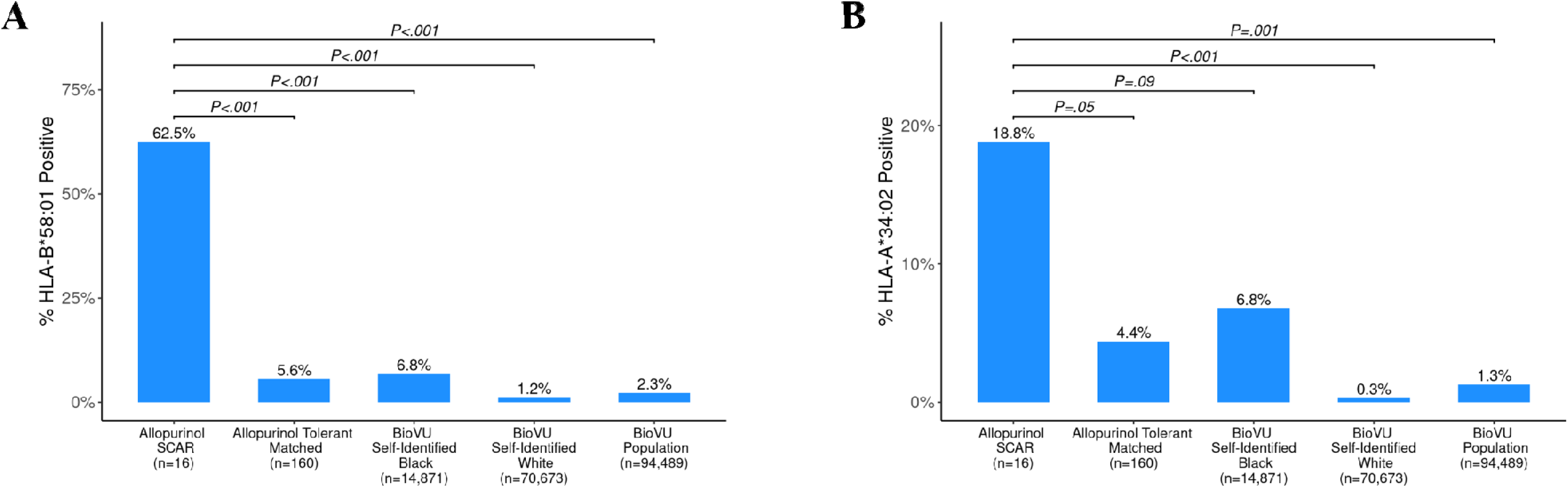
HLA-B*58:01 and HLA-A34:02 carriage are increased in allopurinol-SCAR. **(A**) Percentage of HLA-B*58:01 positive individuals for each described population. (B) Percentage of HLA-A*34:02 positive individuals for each described population.

**Table 2:** HLA-B*58:01 and HLA-A*34:02 are significantly associated with allopurinol-SCAR. Conditional logistic regression analysis for HLA class I to compare allopurinol-SCAR cases and allopurinol tolerant (10:1) matched controls (matched for age, gender, race).

| Allele | Covariates | OR (95% CI) | *P* | *Pc* Bonferroni |
| --- | --- | --- | --- | --- |
| B*58:01 |  | 28 (8.6, 100.6) | <.001 | <.001 |
| A*34:02 | B*58:01 | 20.6 (3.3, 131.1) | <.001 | .04 |
| B*35:01 | B*58:01 + A*34:02 | 10.7 (1.7, 88.7) | .01 | .68 |
| B*58:02 | B*58:01 + A*34:02 + B*35:01 | 32 (1.1, 1054) | .03 | 1 |

## DISCUSSION

We are the first to demonstrate that HLA-B*58:01 is strongly associated with allopurinol-SCAR in a predominantly self-identified US Black population. Homozygosity at HLA-B*58:01 appeared to confer particularly high risk, suggesting a potential gene-dose effect in this population. HLA-A*34:02, an allele associated with allopurinol-induced drug-induced liver injury (DILI), a condition with phenotypic overlap with DRESS^14^, emerged as an additional risk factor for allopurinol-SCAR. HLA-B*35:01, an allele with no previous linkage to allopurinol-SCAR, also appeared to be a potential additional risk factor; however future studies are needed to validate this finding. We also observed a single case of allopurinol-SCAR that carried HLA-B*58:02, an allele closely related to HLA-B*58:01 which would not be detected by HLA-B*58:01 specific testing. Importantly, carriage of HLA-B*58:01, HLA-B*58:02, HLA-A*34:02, and HLA-B*35:01 accounted for 100% of allopurinol-SCAR cases in our patient cohort. This finding suggests that multiple allele testing would more accurately identify representative risk in a heterogenous population^15^. Additionally, this study also demonstrated that those of self-identified Black race exhibited more clinical risk factors such as increased baseline creatine values and prevalence of CKD compared to those of self-identified White race, highlighting clinical factors also associated with allopurinol-SCAR.

### Limitations

While the number of allopurinol-SCAR cases in this study is small, this limitation is offset by the clinical relevance of the population studied which highlights the overrepresentation of CKD and HLA-B*58:01 carriage in Black versus White populations in the US. The ability to determine these associations despite the small sample size and rarity of SCAR reflects upon high effects sizes and the notable safety finding that screening for HLA-B*58:01 alone would miss one-third of allopurinol-SCAR cases in US populations.

## CONCLUSIONS

In our primarily self-identified Black US population, we replicated the established association of HLA-B*58:01 and identified HLA-A*34:02 as a novel risk factor for allopurinol-SCAR. HLA-B*58:01 and CKD are both independently associated with increased risk and are overrepresented in Black populations, underscoring the convergence of genetic susceptibility and the impact of social and structural determinants of health. The established risk allele HLA-B*58:01 was absent in over one-third of patients with allopurinol-SCAR, highlighting the need to expand our understanding of additional genetic and host factors that contribute to risk. These findings support the necessity of population-based studies aiming to mitigate harm among those most vulnerable to these severe, life-threatening reactions.

## Author Contributions

All authors had full access to all of the data in the study and take responsibility for the integrity of the data and the accuracy of the data analysis. CNC and MSK contributed equally to the work. *Acquisition of original cohort*: EJP *Concept and design*: CNC, MSK, EJP *Acquisition, analysis, or interpretation of data*: CNC, MSK, AY, EJP *Drafting of the manuscript*: CNC, MSK *Critical review of the manuscript for important intellectual content*: CNC, MSK, EJP *Statistical analysis*: MSK *Obtained funding*: EJP *Administrative, technical, or material support*: AY, EJP *Supervision*: EJP

## Data Sharing Statement

De-identified clinical data and high-resolution HLA sequencing will be made available to all investigators with or without existing support.

## Data Availability

De-identified clinical data and high resolution HLA sequencing will be made available upon request to the corresponding author.

## Acknowledgements

We gratefully thank and acknowledge the community of SJS/TEN and DRESS patients for their participation in this study. We would like to thank the staff of the Vanderbilt Institute for Clinical and Translational Research, supported by NIH UL1TR002243, for maintaining BioVU and Synthetic Derivative from which the allopurinol tolerant and population controls were derived. We acknowledge the Immunogenetic Microbial Genomics and Single Cell Technologies Core (IMGSCT) at VUMC and the Institute for Immunology and Infectious Diseases (A. Chopra), Murdoch University, Murdoch Western Australia for the provision of class I and II HLA typing. **SJS Survivor Study: Elizabeth J Phillips, MD (PI),** Roni Dodiuk Gad, MD, Aaron Drucker, MD, Elizabeth Ergen, MD, Michelle Goh MBBS, Benjamin Kaffenberger, MD, Robert Micheletti, MD, Rama Gangula, MS, Rebecca Lee, Dana King, Kelby Mahan, Michelle Martin-Pozo, PhD, April O’Connor, Amy Palubinksy, PhD, Suman Pakala, ME, Kristina Williams, Elizabeth Williams, MPH

## FIGURE LEGENDS

**eFigure 1.**
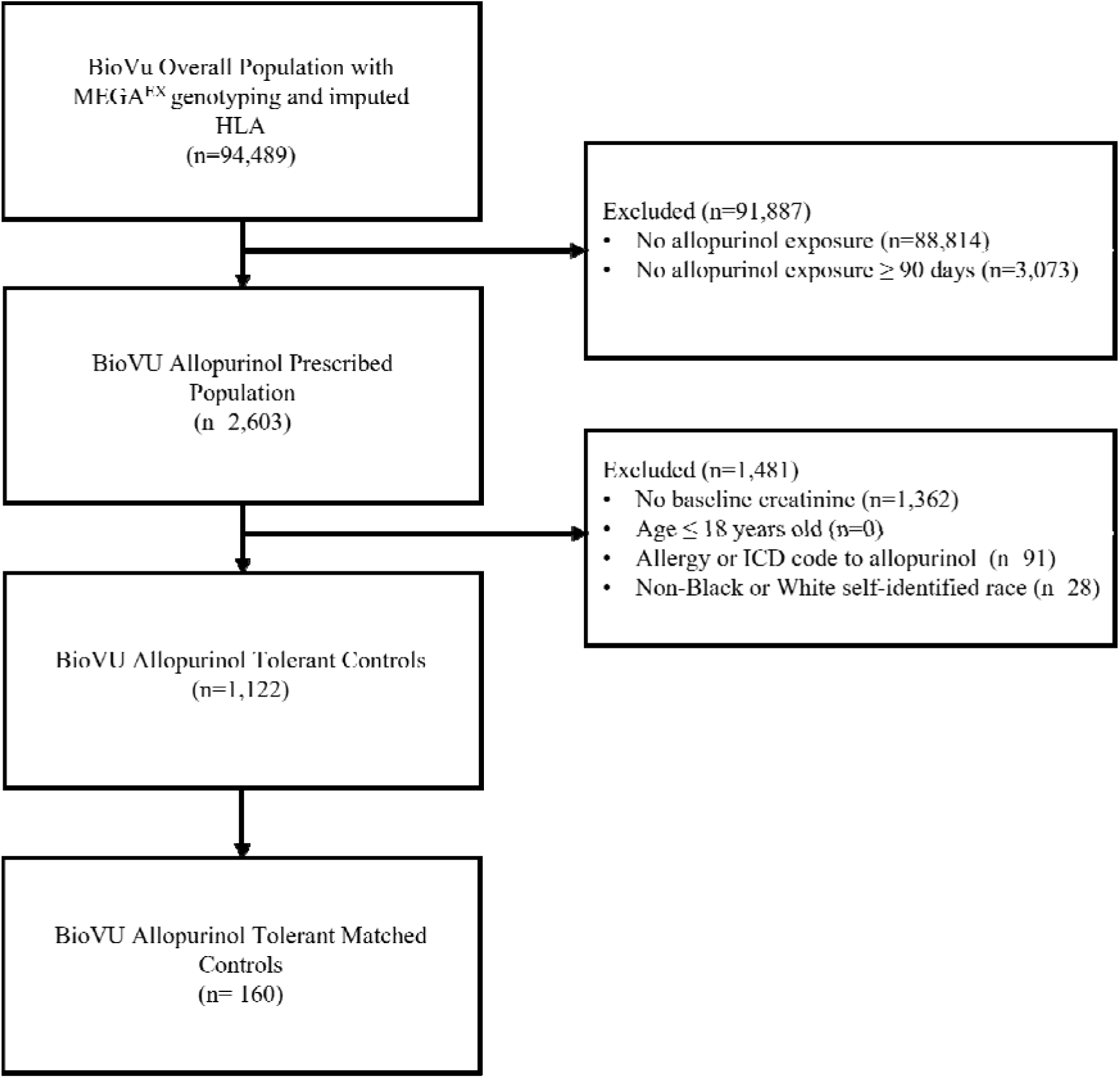
Consort diagram illustrating inclusion criteria for the BioVU overall population and allopurinol tolerant controls.

**eFigure 2:**
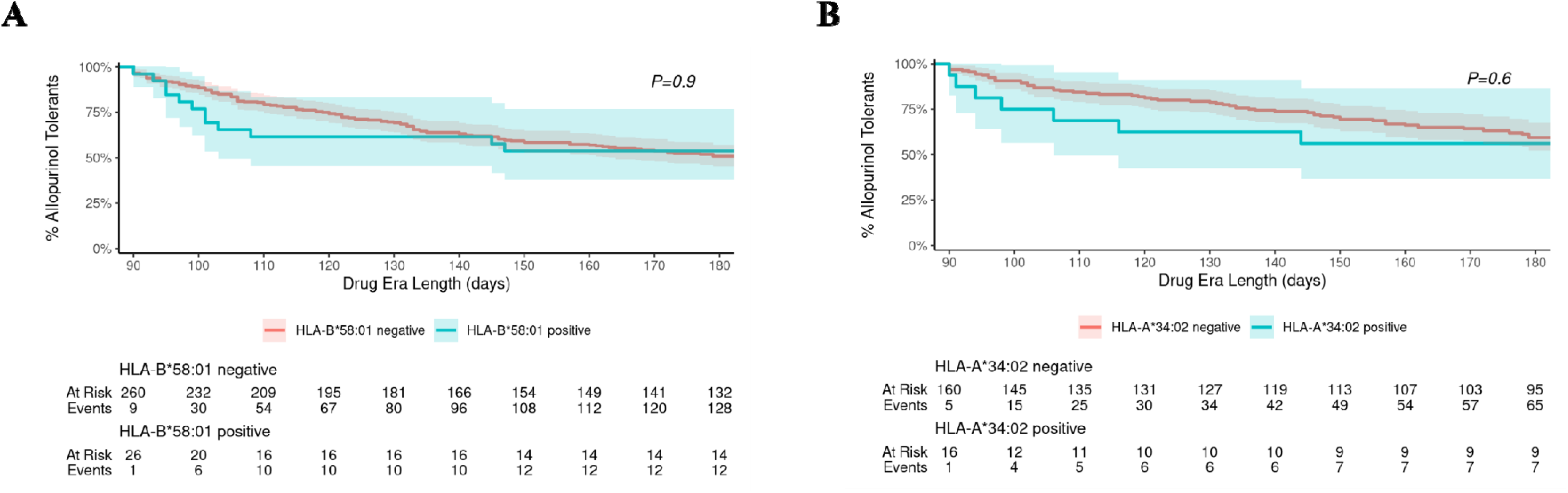
There is no difference in drug era length for allopurinol tolerant controls carrying HLA-B*58:01 or HLA-A*34:02 compared to allopurinol tolerant controls that do not carry the respective HLA alleles. (A) Survival analysis for a drug era length of 180 days comparing allopurinol tolerant controls that are either HLA-B*58:01 positive or negative. (B) Survival analysis for a drug era length of 180 days comparing allopurinol tolerant controls that are either HLA-A*34:02 positive or negative.

**eTable 1:** Complete HLA carriage of the allopurinol-SCAR cases cohort. High-resolution HLA typing was performed by Illumina MiSeq as previously published by sequencing of the Exon 2,3 region of HLA Class I and HLA-DQB1 and the Exon 2 region of HLA-DRB1, DQA1, and DPB1. Reference sequences were obtained through the IMGT/HLA Sequence Database and alleles were called through IIID HLA Analysis Suite. *NP,* allele not present.

|  | HLA-A |  | HLA-B |  | HLA-C |  | HLA-DPB1 |  | HLA-DQA1 |  | HLA-DQB1 |  | HLA-DRB1 |  | HLA-DRB3 |  | HLA-DRB4 |  | HLA-DRB5 |  |
| --- | --- | --- | --- | --- | --- | --- | --- | --- | --- | --- | --- | --- | --- | --- | --- | --- | --- | --- | --- | --- |
| ID | Allele 1 | Allele 2 | Allele 1 | Allele 2 | Allele 1 | Allele 2 | Allele 1 | Allele 2 | Allele 1 | Allele 2 | Allele 1 | Allele 2 | Allele 1 | Allele 2 | Allele 1 | Allele 2 | Allele 1 | Allele 2 | Allele 1 | Allele 2 |
| 1 | 29:02 | 68:02 | 15:03 | 58:01 | 2:10 | 07:01 | 01:01 | 11:01 | 01:02 | 02:01 | 02:02 | 06:02 | 07:01 | 15:03 | NP | NP | 01:01 | NP | 01:01 | NP |
| 2 | 02:02 | 02:02 | 58:01 | 58:01 | 08:02 | 08:02 | 01:01 | 04:02 | 01:01 | 03:01 | 03:03 | 05:01 | 13:01 | 14:01 | 02:02 | 02:02 | NP | NP | NP | NP |
| 3 | 23:01 | 30:01 | 42:01 | 58:01 | 07:01 | 17:01 | 01:01 | 01:01 | 04:01 | 05:01 | 02:01 | 04:02 | 03:01 | 03:01 | 01:01 | 02:02 | NP | NP | NP | NP |
| 4 | 29:02 | 74:01 | 35:01 | 53:01 | 04:01 | 07:01 | 02:01 | 17:01 | 01:02 | 04:01 | 03:01 | 05:01 | 08:04 | 13:02 | 03:01 | NP | NP | NP | NP | NP |
| 5 | 03:01 | 23:01 | 49:01 | 58:02 | 04:01 | 07:01 | 01:01 | 01:01 | 03:01 | 03:01 | 02:02 | 03:02 | 04:05 | 07:01 | NP | NP | 01:01 | 01:01 | NP | NP |
| 6 | 01:01 | 26:01 | 08:01 | 58:01 | 07:01 | 07:01 | 03:01 | 04:01 | 03:01 | 04:01 | 03:02 | 04:02 | 04:04 | 08:04 | NP | NP | 01:01 | NP | NP | NP |
| 7 | 03:01 | 25:01 | 07:02 | 58:01 | 03:02 | 07:02 | 04:01 | 14:01 | 01:02 | 01:02 | 06:02 | 06:09 | 13:02 | 15:01 | 03:01 | NP | NP | NP | 01:01 | NP |
| 8 | 30:01 | 30:02 | 35:01 | 42:01 | 04:01 | 17:01 | 02:01 | 04:02 | 02:01 | 04:01 | 02:02 | 04:02 | 03:02 | 07:01 | 01:01 | NP | 01:01 | NP | NP | NP |
| 9 | 23:01 | 34:02 | 35:01 | 44:02 | 04:01 | 05:01 | 04:01 | 04:02 | 01:02 | 03:01 | 03:01 | 06:02 | 04:01 | 11:01 | 02:02 | NP | 01:01 | NP | NP | NP |
| 10 | 23:01 | 74:01 | 58:01 | 58:01 | 03:02 | 07:01 | 01:01 | 18:01 | 01:02 | 02:01 | 02:02 | 06:02 | 07:01 | 11:01 | 02:02 | NP | 01:01 | NP | NP | NP |
| 11 | 30:02 | 36:01 | 13:02 | 58:01 | 08:04 | 16:01 | 02:01 | 13:01 | 01:02 | 04:01 | 04:02 | 06:02 | 08:04 | 15:03 | NP | NP | NP | NP | 01:01 | NP |
| 12 | 01:01 | 34:02 | 14:02 | 35:01 | 04:01 | 08:02 | 04:02 | 04:02 | 01:01 | 01:01 | 05:01 | 05:01 | 01:01 | 01:02 | NP | NP | NP | NP | NP | NP |
| 13 | 11:01 | 25:01 | 18:01 | 58:01 | 05:01 | 07:01 | 04:01 | 04:01 | 01:02 | 03:01 | 03:02 | 06:02 | 04:01 | 15:01 | NP | NP | 01:01 | NP | 01:01 | NP |
| 14 | 02:05 | 03:01 | 13:02 | 58:01 | 06:02 | 07:01 | 04:01 | 17:01 | 02:01 | 02:01 | 02:02 | 02:02 | 07:01 | 07:01 | NP | NP | 01:01 | 01:01 | NP | NP |
| 15 | 23:01 | 33:03 | 15:16 | 58:01 | 07:01 | 14:02 | 01:01 | 17:01 | 02:01 | 02:01 | 02:02 | 02:02 | 07:01 | 07:01 | NP | NP | 01:01 | 01:01 | NP | NP |
| 16 | 23:01 | 34:02 | 14:03 | 44:03 | 07:01 | 08:02 | 02:01 | 03:01 | 01:01 | 01:02 | 05:01 | 06:09 | 10:01 | 13:02 | 03:01 | NP | NP | NP | NP | NP |

**eTable 2:** Characteristic summary of the BioVU allopurinol tolerant control cohort according to self-identified race.

| **Variable** | **Black**<br>**N = 212<sup>1</sup>** | **White**<br>**N = 910<sup>1</sup>** | **P<sup>2</sup>** |
| --- | --- | --- | --- |
| **Age** | 63 (54, 71) | 66 (58, 73) | .004 |
| **Gender** |  |  | .004 |
| Female | 82 (39%) | 260 (29%) |  |
| Male | 130 (61%) | 650 (71%) |  |
| **Drug Era Length** | 228 (122, 1,752) | 177 (115, 704) | .04 |
| **Creatinine** | 1.76 (1.38, 2.83) | 1.46 (1.16, 2.00) | <.001 |
| **HLA-B\*58:01 Positive** | 12 (5.7%) | 14 (1.5%) | .001 |
| **HLA-A\*34:02 Positive** | 14 (6.6%) | 2 (0.2%) | <.001 |
| **CKD** | 191 (90%) | 670 (74%) | <.001 |
| **Gout** | 195 (92%) | 781 (86%) | .02 |
| **CKD and Gout** | 177 (83%) | 610 (67%) | <.001 |

<sup>1</sup>Median (Q1, Q3); n (%), <sup>2</sup>Wilcoxon rank sum test; Pearson’s Chi-squared test; Fisher’s exact test

**eTable 3:** Homozygosity at HLA-B*58:01 increases risk of allopurinol-SCAR. Fisher’s exact test and calculated OR and 95% CI with Haldane’s modification for heterozygosity and homozygosity at HLA-B*58:01 to evaluate gene-dose effect.

| Genotype | OR (95% CI) | *P* |
| --- | --- | --- |
| B*58:01 Heterozygous | 28 (8.6, 100.6) | <.001 |
| B*58:01 Homozygous | 55.3 (2.5, 1208.6) | .01 |

